# Epidemiological transition in Australia: An analysis of immigration patterns in relation to circulatory system diseases and all-cause mortality in the mid-twentieth century

**DOI:** 10.1101/2022.12.11.22282585

**Authors:** Cecily C Kelleher, Gabrielle E Kelly, Ricardo Segurado, Jonathan Briody, Alexander M Sellers, Janet S McCalman

## Abstract

**Background and Objectives:** Circulatory System Diseases (CSD) patterns vary over time and between countries, related to lifestyle risk factors, associated in turn with socio-economic circumstances. Current global CSD epidemics in developing economies are similar in scale to those observed previously in the USA and Australasia. Australia exhibits an important macroeconomic phenomenon as a rapidly transitioning economy with high immigration throughout the 19^th^ and 20^th^ centuries. We wished to examine how that historical immigration related to CSD patterns subsequently.

**Methods and Setting:** We provide a novel empirical analysis employing census-derived place of birth by age bracket and sex from 1891 to 1986, in order to map patterns of immigration against CSD mortality rates from 1907 onwards. Age-specific generalised additive models for both CSD mortality in the general population, and all-cause mortality for the foreign-born (FB) only, from 1910 to 1980 were also devised for both males and females.

**Results:** The % FB fell from 32% in 1891 to 9.8% in 1947. Rates of CSD rose consistently, particularly from the 1940s onwards, peaked in the 1960s, then declined sharply in the 1980s and showed a strong period effect across age groups and genders. The main effects of age and census year and their interaction were highly statistically significant for CSD mortality for males (p < 0.001, each term) and for females (p < 0.001, each term). The main effect of age was statistically significant for all-cause mortality minus net migration rates for the FB males (p =0.005) and for FB females, both age (p < 0.001) and the interaction term (p=0.002) were significant.

**Conclusions:** We argue our empirical calculations, supported by historical and socio-epidemiological evidence, employing immigration patterns as a proxy for epidemiological transition, affirm the lifecourse hypothesis that both early life circumstances and later life lifestyle drive CSD patterns.

**Article Summary:** *Strengths:*

- An original analysis employing census data and immigration patterns to reinterpret historical trends in CSD in Australia
- Relevant to modern public health policy for population approaches to CSD prevention, also integrates lifecourse and lifestyle drivers of trends

*Limitations:*

- Historical databases do not categorise either all cause or CSD mortality according to country of origin.
- However, data for foreign-born mortality were inferred using novel actuarial type calculations
- There are no second-generation data by country of origin, unlike in USA.

## Introduction

Circulatory system diseases (CSD) patterns are known to vary over time and between countries, related to lifestyle risk factors across the life course, associated in turn with underlying socio-economic circumstances^1-6^. Current global epidemics in developing economies are similar in scale to those observed much earlier in the USA and Australia^7-8^. Since the mid twentieth century in particular, international epidemiological studies have assessed how proximal lifestyle risk factors such as tobacco smoking and diet-related hypertension, obesity and lipoprotein profiles can explain the patterns of heart disease and stroke in given populations^7-13^.

It is well established that in the mid-twentieth century, patterns of heart disease were highest in countries such as the USA, Australia and Finland, intermediate in northern Europe and lowest in Mediterranean countries^14^. That pattern shifted, with steep declines observed first in the USA and Australia, followed by falls in all western countries from the early 1980s onwards^14-15^. The conventional, primarily period, explanation is that the balance of lifestyle change and introduction of rigorous clinical management protocols, including coronary care and pharmacological management of hypertension and hypercholesterolemia drive these patterns^5, 7, 11, 16^. The IMPACT models suggest that declining incidence is driven primarily by shifts in risk factors, rather than treatments^4^. However, the “causes of causes” hypothesis suggests that these lifestyle patterns are strongly influenced by socio-economic forces and that in turn a life course perspective is required to understand how cumulative, or trajectory influences occur over time, suggesting cohort variations^17^.

The concept of epidemiological transition posits that as countries move from patterns of poverty to patterns of affluence, changes in prevailing diseases occur, with first infectious diseases waning, followed by a rise in non-communicable diseases^1,18,19^. The American Heart Disease Association describes four distinct stages of epidemiological transition at population level, stage 1 being pestilence and famine associated with heart conditions including rheumatic heart disease, stage 2 being receding pandemics which also now include a rise in hypertensive heart disease and haemorrhagic stroke, stage 3 being degenerative and man-made diseases with all forms of strokes, ischaemic heart disease at all ages and increasing obesity and diabetes and stage 4 is associated with delayed degenerative diseases with onset of stroke, ischaemic heart disease and heart failure at older ages^1^.

Harper et al. set out a contextual framework for understanding these trends which takes account of socio-economic position of individuals, lifecourse processes and the macrosocial context^14^. The contemporary global burden of disease maps show clearly the shifting patterns downwards in current Western populations and upwards in parts of Asia and Africa^20^. Barker observed the paradox that while coronary heart disease increases with western prosperity, it is more common in poorer areas of western countries and among poorer peoples^21.^ Moreover, in a time of huge global challenge in relation to non-communicable disease and the rise of obesity in recent decades, it is necessary to understand at a mechanistic level how global epidemics occur to intervene effectively in their management^22, 23, 24^. An assessment of how historical patterns of disease occurred, can contribute to modern strategies to prevent non communicable disease^6^.

The USA, Australia and New Zealand share an important macroeconomic phenomenon in that they were each rapidly developing transitional economies throughout the nineteenth and twentieth centuries, with large scale immigration as a major driver of population growth and composition^15, 25-27^. In both continents the original immigrants were predominantly from Europe, initially from Northern and then Central Europe, followed by Southern Europe and then latterly from Asia.

An analysis employing historical USA census data, showed a strong temporal association between the patterns of age-adjusted heart disease in both men and women and the proportion of foreign born and of first-generation Americans between thirty-eight and fifty years earlier^15^. This was possible because the demographic changes occurring on such a large scale meant that the census recorded country of birth of enumerated householders^8, 26, 27^. Stamler suggested that the foreign-born might exhibit residual features reflecting the influence of their country of origin, i.e., a cohort effect, but their characteristics as Americans were predominant^8^.

## Methods

### Data

To examine the transitioning population and its subsequent pattern of CSD we looked at both census data and epidemiological records and constructed a historical dataset based on digitised paper records. Census data on place of birth, age bracket, and sex were retrieved from scanned paper copies from 1891 over the subsequent 95 years coming to thirteen in total – every 10 years between 1891 and 1921, 1933, 1947, 1954, and then every 5 years from 1961 to 1986^28^. Prior to 1891 there was not a synchronised census year for all six pre-Federation colonies, however an overall number of foreign-born residents were obtained for 1861, 1871 and 1881 (not by region).

We reconciled the country of birth data across changes of political entities into: Commonwealth of Australia (the six colonies/states, External Territories, and New Zealand), UK and Ireland, Northern and Central Europe (including Denmark, Sweden and Norway, Germany, The Netherlands, Belgium, France, Switzerland, Austria/Austria-Hungary, Eastern Europe through to Russia), Southern Europe (all other countries with extensive Mediterranean sea coasts including Spain, Italy, Greece, Malta, Yugoslavia and other Balkan nations, Turkey, and Lebanon, in addition to Portugal, Romania and Bulgaria), with the Americas, Asia, Africa, and Polynesia grouped into the Other group including the small number of persons unknown, or born at sea. The percentage of the total population which were born in each of the world regions was calculated. As the predominant immigrant population of Australia historically has been from the UK and Ireland, we also created an aggregated European and Other region, to include all non-UK/Irish immigration.

Age-specific and age-adjusted circulatory system disease (CSD) mortality rates per 100,000 were available from 1907 through to 2016 from online records of the General Record of Incidence of Mortality (GRIM; Australian Institute of Health and Welfare) book 0900^29^. The GRIM mortality information is obtained from (compulsory) death certificates, certified by a medical practitioner or coroner, and coded by the Australian Bureau of Statistics or precursors to that agency, to International Statistical Classification of Diseases and Related Health Problems (ICD). CSD currently encompasses ICD-10 codes I00-I99 and is aligned with corresponding categories back to ICD-1 (1907-1917). Coronary heart disease deaths specifically (ICD-10 codes I20-I25) were considered but were only available commencing in 1940. We obtained incidence of aggregated cerebrovascular disease (CVD) mortality from GRIM book 0904 (Australian Institute of Health and Welfare), encompassing ICD-10 codes I60 to I69. All mortality rates are age-standardised to the 2001 estimated resident Australian population.

### Analysis

Age-specific mortality rates for the whole Australian population are also compared with those foreign-born^30,31^. CSD mortality rates are available only for the whole Australian population but there is justification that these are indicative of all-cause mortality rates. For each sex/age-group/census year mortality rates due to CSD in the Australian population were calculated by taking the ratio N_mort/N_total in the raw data set.

Mortality rates for foreign born due to circulatory disease or all-causes are not readily available either, as country of origin was not recorded at death. However, all-cause mortality rates for foreign born were estimated from numbers of foreign born in different age groups at different census years, using the following actuarial type/analytical method. For example, let fb denote foreign born, and taking a specific age and specific year, then Number fb age 31 in 1948 = (Number fb age 30 in 1947) × (1-(r_1_-r_2_)), where r_1_ is the mortality rate for fb age 30 1947and r_2_ is the inward migration rate for age 30 in 1947-1948. Let r=r_1_-r_2_ then we re-write the above equation as:

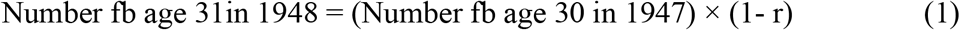

Thus, r is the mortality rate-inward migration rate for the fb age 30 in 1947. Then

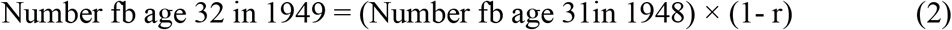

where r=r_1_-r_2_ and r_1_ is the mortality rate for fb age 31 1948-1949 and r_2_ is the inward migration rate for age 31 in 1948-1949 and we assume these rates are unchanged from the 1947-1948 values.

Thus, substituting equation (1) into equation (2), we have

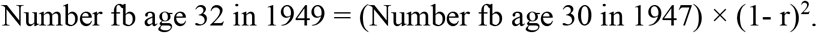

Proceeding in this way we have

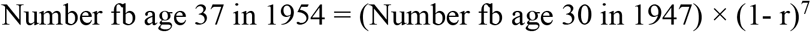

Therefore

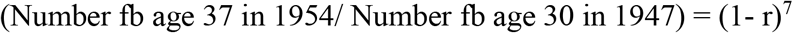

and

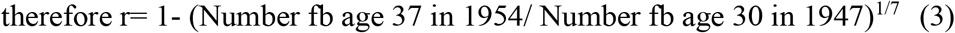

Thus, r is the mortality rate-inward migration rate for the fb age between 30 and 36 per year in the interval 1947-1954.

r is estimated from the data by

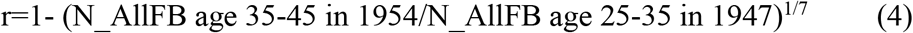

This calculation was repeated for each age-group and census interval. Only raw data by age-group totals are available from the censuses. Note that r can be negative if the inward migration rate exceeds the mortality rate. Thus, for all age-groups r will underestimate the mortality rates but it is used here as a proxy for the mortality rates.

The estimates of r were interpolated and smoothed over all ages and census years using a tensor product smooth model^32^ and are displayed in 3-d plots. This is an extension of a generalized additive model (gam)^33^ to 3-dimensions and was fitted using the mgcv package in R with the ti function. This was done separately for males and females. This smooth model was decomposed into a functional ANOVA structure with main effects of age and year and their interaction.

The CSD mortality rates per 100,000 for the whole population were also fitted using a tensor smooth model and displayed in 3-d plots.

Note that the raw data are very variable both by census year and age-group. For example, the number of foreign-born males age group 30 in 1921 is 74,913 while it is 100,385 in 1933 and for age group 40 in 1921 it is 93,089. So, all interpolating models must be interpreted with caution.

## Results

The exact population numbers from 1891-1986 from census data by region of origin are presented in Table 1. This shows respectively the population numbers born in Australia or abroad or the birthplace of origin as a percentage of the total population, from 1891 onwards. There are two distinct patterns observed. In 1891 when the population numbered just over three million, the proportion foreign-born was relatively high. The contribution made to the population composition by those UK or Irish-born is initially very considerable but tails off until the 1940s. A new surge in migration beginning after WW2 is apparent, initially comprising an increase in European (mostly Southern) immigration. Asian populations in particular were the predominant group in the large “Other” category after the 1970s.

**Table 1.** Descriptive statistics of Australian population composition by country of birth, from census data 1891 - 1986.

| Year | Total | UK/Ireland |  | Northern Europe |  | Southern Europe |  | Other Place of origin |  | All Foreign Born |  | Australian Born |  |
| --- | --- | --- | --- | --- | --- | --- | --- | --- | --- | --- | --- | --- | --- |
|  |  | N | % | N | % | N | % | N | % | N | % | N | % |
| 1891 | 3,174,392 | 821,166 | 26% | 75,145 | 2.4% | 5,307 | 0.2% | 113,799 | 4% | 1,015,417 | 32.0% | 2,158,975 | 68.0% |
| 1901 | 3,773,801 | 679,159 | 18% | 67,291 | 2% | 7,382 | 0% | 111,666 | 3% | 865,498 | 23% | 2,908,303 | 77.0% |
| 1911 | 4,455,005 | 591,729 | 13% | 63,594 | 1.4% | 9,348 | 0.2% | 122,664 | 3% | 787,335 | 17.7% | 3,667,670 | 82.3% |
| 1921 | 5,435,210 | 676,387 | 12% | 52,503 | 1.0% | 15,539 | 0.3% | 109,118 | 2% | 853,547 | 15.7% | 4,581,663 | 84.3% |
| 1933 | 6,646,905 | 732,674 | 11% | 48,125 | 0.7% | 44,680 | 0.7% | 94,860 | 1% | 920,339 | 13.8% | 5,726,566 | 86.2% |
| 1947 | 7,577,772 | 542,910 | 7% | 50,180 | 0.7% | 58,516 | 0.8% | 90,995 | 1% | 742,601 | 9.8% | 6,835,171 | 90.2% |
| 1954 | 8,894,884 | 664,205 | 7% | 294,667 | 3.3% | 196,192 | 2.2% | 39,756 | 0% | 1,194,820 | 13.4% | 7,700,064 | 86.6% |
| 1961 | 10,504,210 | 755,402 | 7% | 433,999 | 4.1% | 406,791 | 3.9% | 178,612 | 2% | 1,774,804 | 16.9% | 8,729,406 | 83.1% |
| 1966 | 11,550,462 | 908,664 | 8% | 430,574 | 3.7% | 554,273 | 4.8% | 237,409 | 2% | 2,130,920 | 18.4% | 9,419,542 | 81.6% |
| 1971 | 12,755,638 | 1,088,210 | 9% | 475,095 | 3.7% | 633,173 | 5.0% | 382,840 | 3% | 2,579,318 | 20.2% | 10,176,320 | 79.8% |
| 1976 | 13,548,649 | 1,117,600 | 8% | 460,673 | 3.4% | 632,550 | 4.7% | 508,209 | 4% | 2,719,032 | 20.1% | 10,829,617 | 79.9% |
| 1981 | 14,576,330 | 1,132,599 | 8% | 471,278 | 3.2% | 678,445 | 4.7% | 900,143 | 6% | 3,182,465 | 21.8% | 11,393,865 | 78.2% |
| 1986 | 15,602,150 | 1,127,191 | 7% | 488,825 | 3.1% | 662,124 | 4.2% | 1,213,554 | 8% | 3,491,694 | 22.4% | 12,110,456 | 77.6% |

The Australian population composition mapped against the age-adjusted circulatory system disease (CSD) mortality is shown in Figure 1a. This illustrates the change in the proportions of the population foreign-born by region of origin (left-hand vertical axis), alongside the change in incident mortality due to all CSD (red line, right-hand vertical axis). This shows firstly that the percentage of the population foreign-born was over 60% in 1861 and declined steeply over the next 80 years so that it was just 10% by the mid nineteen forties. Thereafter there was again a rise in immigration, climbing to over 20% of the population in the early 1980s. The figure also shows the contribution to this pattern made by the UK/Ireland, Northern and Southern European groups. The predominant early group is of UK and Irish origin, dropping steeply by the mid nineteen forties and plateauing at approximately 7%. The other two European groups are much smaller in proportion and reach their peak in the 1960s.

**Figure 1a:**
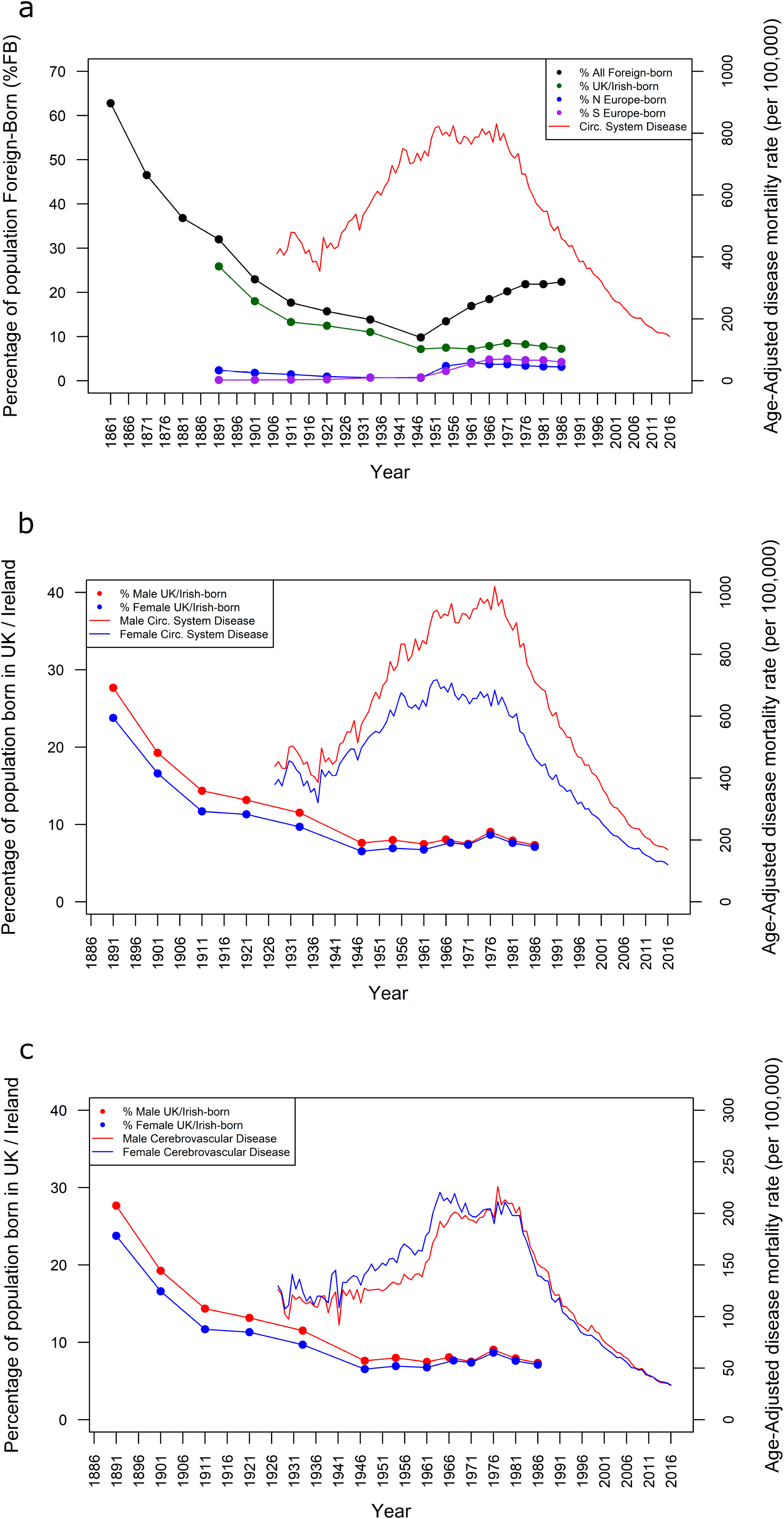
Timeline of foreign-born census respondents 1861-1986 and age-adjusted circulatory system disease mortality for the Total Australian population Figure 1b: Sex-specific Irish/British-born census respondents and age-adjusted circulatory system disease mortality for both males and females in the Australian population Figure 1c: Sex-specific Irish/British-born census respondents against age-adjusted cerebrovascular disease mortality for the Total Australian population

The pattern of circulatory diseases shows a steep rise throughout the early twentieth century, and reaches a plateau by 1950-1970, and then declines sharply from then onwards. By 1990 rates had fallen again to those previously seen in 1920 and continued to decline before levelling off currently.

Figure 1b shows the contrast between males and females in age-adjusted CSD mortality, this time in relation to the population foreign born of both sexes from UK and Ireland. There is an approximate time lag of forty years in the fall-off of immigration from UK and Ireland and that of age adjusted CSD in the general population. Female and male rates began to diverge in the 1940-50s, with female rates declining earlier, and male rates not beginning to decline until the 1980s. By the late 1990s the rates had converged again. The pattern in Figure 1c showing the cerebrovascular disease mortality by sex, aligns with this, albeit with negligible sex differences: an increase as the late 19^th^ century immigrants reach old age, a plateau from the 1950s to 1970s, followed by steep decline thereafter.

Figures 2a and 2b show mortality rates due to circulatory disease in the Australian population by age group and year during the period 1910 to 1990 for males and females separately. Mortality rates increase with age as expected and can be seen from the left scales. In all age groups and in both males and females, rates of circulatory disease rose consistently, particularly from the 1940s onwards, peaked in the late nineteen sixties and then began to decline sharply by the nineteen eighties. An interaction effect between age and year is clearly visible, with differences between ages changing with year. The main effects of age and census year and their interaction were statistically significant in the model for males (p < 0.001, each term) and that for females (p < 0.001, each term). The adjusted R^2^ was 96.4% for the male model and 98% for the female model.

**Figure 2.**
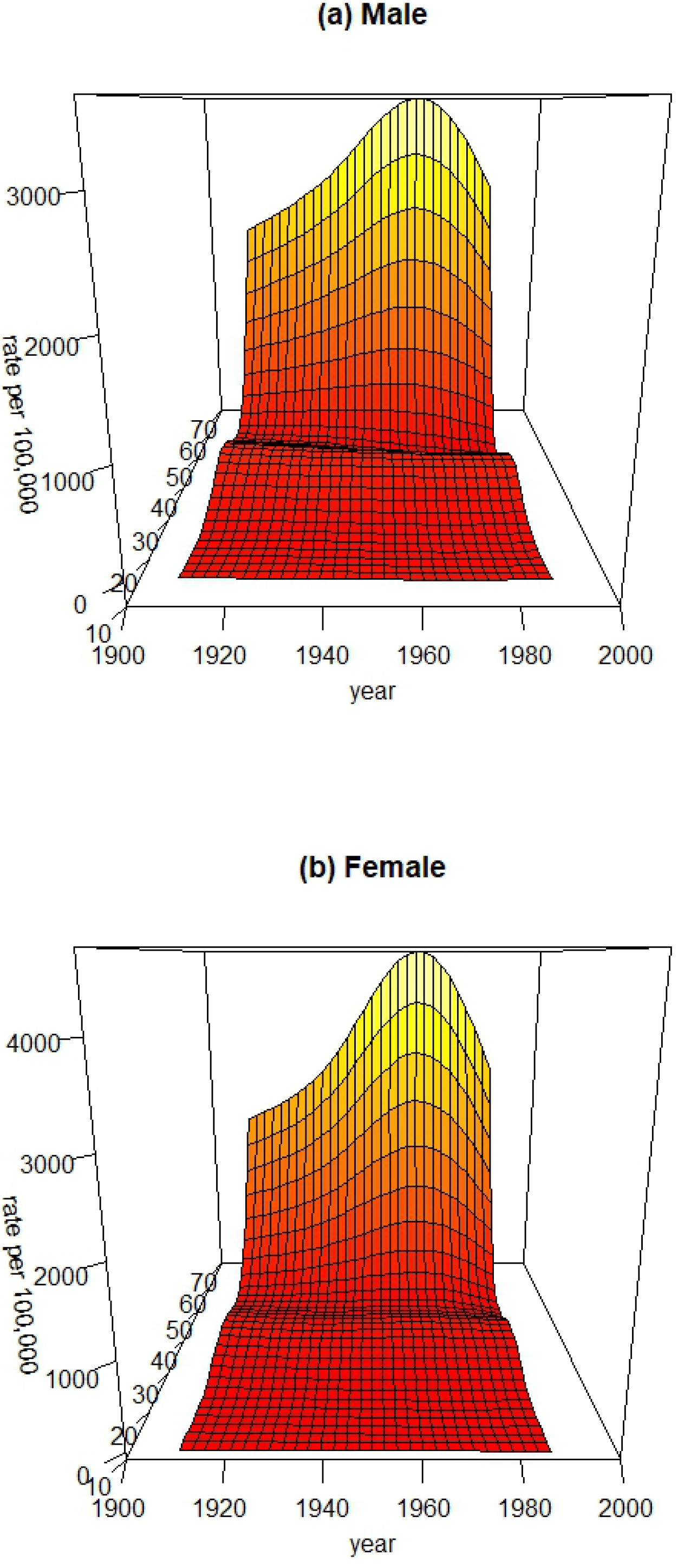
Circulatory system mortality rates per 100,000 by age and year in the general Australian population (a) Males (b) Females. The unlabelled axis is age in both plots.

Figures 3a and 3b show all-cause mortality-net migration rates for the foreign born only, separately for both males and females. The main effect of age was statistically significant in the model for males (p =0.005) and for females both age (p < 0.001) and the interaction term (p=0.002) were significant. The adjusted R^2^ was 35.1% for the male model and 53% for the female model.

**Figure 3.**
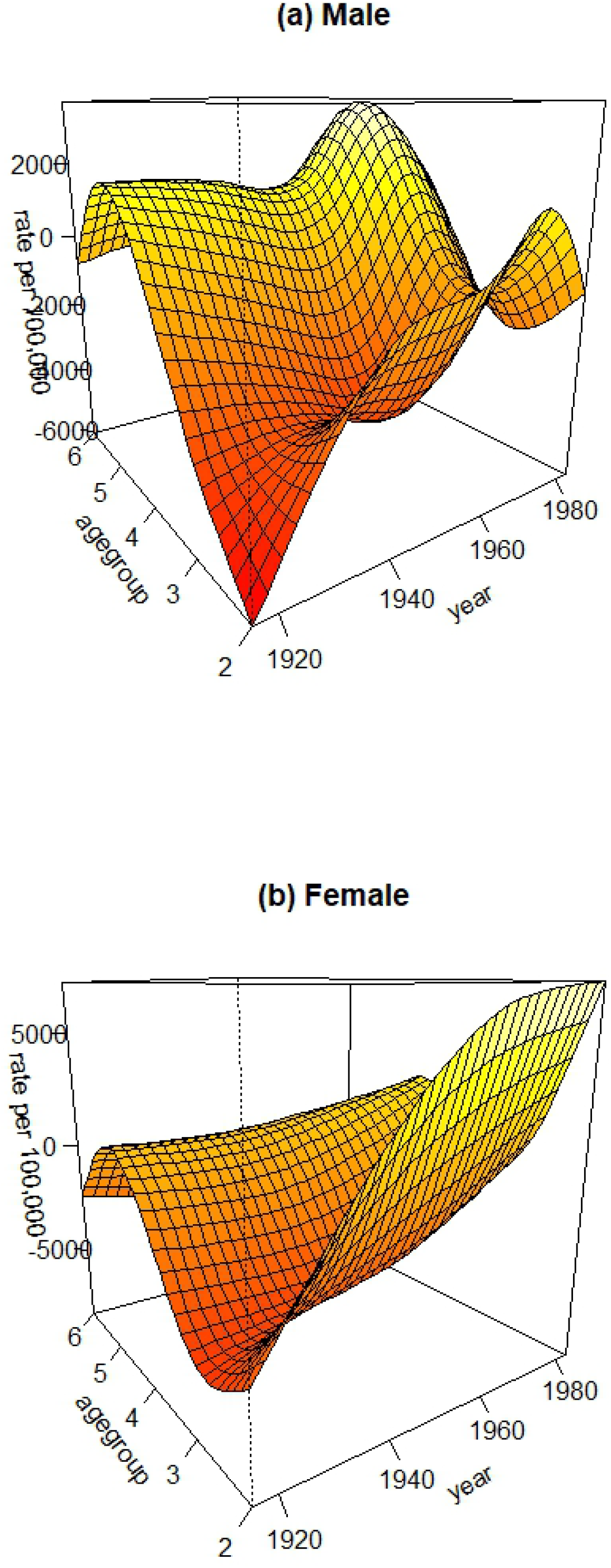
All-cause mortality-net migration rates per 100,000 by age group and year for Australian foreign-born population (a) Males (b) Females. Age groups 2-6 correspond to age midpoints 20,30,40,50,60 years old respectively. Both plots have been rotated with an angle of −35° to permit a better view.

## Discussion

In this analysis we have examined whether the immigration pattern in Australia, as one form of proxy for epidemiological transition, related to patterns of both CSD and all-cause mortality in the population, as had been observed previously in the USA^15^. We demonstrate a temporal association of age-adjusted CSD with immigration approximately a half century earlier from predominantly United Kingdom and Ireland, which changes as the population stabilises economically and newer waves of immigration from lower risk Mediterranean and later Asian countries occur.

We also show in age-specific models that assess both CSD and all-cause mortality, a strong age and period effect for the general Australian population and a highly significant interaction between age and census year in CSD mortality. Both foreign born male and female populations showed a strong age effect for all-cause mortality, while for females the interaction term was also highly significant. These novel findings do not contradict the period impact on cardiovascular disease shown in the age-period-cohort analysis by Taylor et al^5^ for the general Australian population but is unlike their analysis in showing a clear interaction effect, and for the first time looks specifically also at the foreign-born population.

We demonstrate in essence, for Australia, similar temporal findings to those observed in the USA. In the previous analysis, data were available both for foreign born and native born of foreign parents and that afforded a fit which explained both the rise and fall of the epidemic^15^, though we do not have second generation data in this analysis. We acknowledge there are clear limitations to this current analysis in that the historical databases do not categorise either all cause or CSD mortality according to country of origin and there are no available second-generation data by country of origin. However, data for foreign-born were inferred using actuarial type calculations and a three-dimensional model, representing a novel approach. One would expect an impact on foreign-born, but also their children born into a transitioning economic environment, especially urban disadvantage.

Our findings can be situated in the earlier history of that transitioning Australian development. To understand any cohort effect, we contend the historical developments influencing early childhood must be considered. Of the three classical risk factors, active individual smoking is strongly period driven ^7,9^, whereas dietary intake, which drives both lipoprotein and blood pressure patterns, can be influenced both by short term and longer-term programming factors ^6,23,24^.

### Demographic Transition

Borrie provided a comprehensive demographic analysis of the transitioning Australian and New Zealand populations over a two-hundred-year period from 1788-1988^34^. Those who migrated into Australia from Europe^35-37^ included First Fleeter and other colonists, transported convicts, and Assisted Passage individuals who were supported to emigrate from their countries of origin. The Irish and British were the predominant early migrating group. A total of over 800,000 immigrants came also through Government Assistance programmes established 1861-65 through to 1900, tapering off sharply during that time. Irish assisted emigration to Australia was related to workhouse programmes established during and after the Great Irish famine^36-37^. The impact of historical circumstances, including the famine in Ireland in the 1840s has been examined previously in an Irish context, both at home and in relation to its huge emigrant diaspora^38^. Irish emigrants in the United Kingdom took at least two generations to reflect integrated patterns of mortality of the prevailing population^39^. An analysis of birthplace of all brides marrying in Australia between 1908 and 1979 shows the transition to Australian born mothers almost fully completed by 1979^34^.

Whitewell et al conducted a detailed economic and public health review of Australia from 1860-1940 which illustrates how and when population changes occurred which might have influenced longer term health outcomes into the twentieth century^40^. Height, as a proxy measure for public health impact of standards of living, was initially high, dipped in the latter period of the nineteenth century, then rose again from the late eighteen nineties through to 1918. From 1890 onwards, the sanitary conditions in the booming cities, which initially made for an unhealthy environment in which children were especially vulnerable, greatly improved. Inwood et al show an association between growing incomes and height in Tasmania related to a decline in food cost and increased per capita domestic product^41^. Cumpston^26^ documented the improvements in public health in Australia, that would translate into reduced susceptibility to strokes and heart disease in these individuals as older adults. Taylor et al reviewed both all-cause mortality in Australia from 1788-1990^42^ and more recent cause-specific mortality from 1907-1990^43^. They showed a precipitous fall in infant mortality from 1900, similar to European countries, but then a stagnation during the mid-twentieth century, attributable to the epidemic in CSD mortality.

There is now a strong body of recently assembled historical retrospective cohort studies that afford an opportunity to review social and economic circumstances in the late nineteenth and early twentieth centuries when Australia was becoming established as a modern economy and when cardiovascular diseases incidence and mortality were rising^44-49^ McCalman et al linked medically validated morbidity and mortality records to original maternity records from the Melbourne lying-in hospital^44-46^ showing expected demographic predictors of infant mortality. These included reduced odds of that outcome if of higher social group, of Australian birth, and higher odds if a first-born child, younger mother, pre-term and of relatively lower birthweight^46^. Early results from the Victoria Diggers to Veterans longitudinal study of first world war enlisters showed a strong social gradient according to military rank for height and amongst survivors to 1922, farmers were significantly less likely to die compared to others, whereas poorer urban counterparts had greater odds of mortality^49^

### Lifestyle Hypothesis

As in the USA, there was considerable research interest in Australia and New Zealand in the aetiology of non-communicable disease from mid-nineteen seventies onwards^50-59^. Seminal epidemiological analyses document the rising rates of smoking in the population and changes in food availability, exposing a transitioning population to prevailing adverse lifestyle ^7, 9^. Those studies at the height of the epidemic focused not primarily on the early British migrants however but on the latterly arrived mainly Southern European immigrants who initially carried the reduced risk associated with their Mediterranean lifestyle into a now economically stable Australian society (Table 2).

**Table 2:**
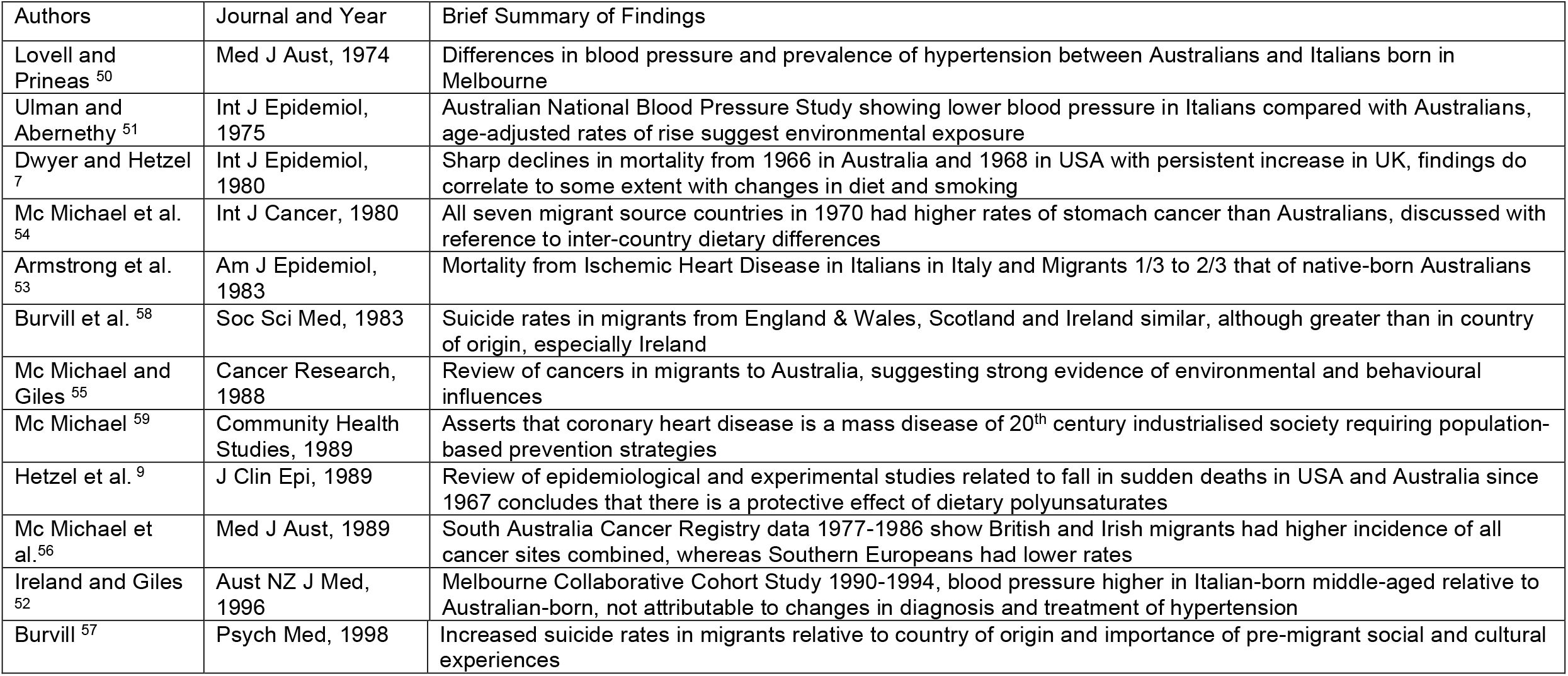
Studies of Lifestyle, Country of Origin and Non-Communicable Disease outcomes in Mid-Twentieth Century Australia.

| Authors | Journal and Year | Brief Summary of Findings |
| --- | --- | --- |
| Lovell and Prineas <sup>50</sup> | Med J Aust, 1974 | Differences in blood pressure and prevalence of hypertension between Australians and Italians born in Melbourne |
| Ulman and Abernethy <sup>51</sup> | Int J Epidemiol, 1975 | Australian National Blood Pressure Study showing lower blood pressure in Italians compared with Australians, age-adjusted rates of rise suggest environmental exposure |
| Dwyer and Hetzel <sup>7</sup> | Int J Epidemiol, 1980 | Sharp declines in mortality from 1966 in Australia and 1968 in USA with persistent increase in UK, findings do correlate to some extent with changes in diet and smoking |
| Mc Michael et al. <sup>54</sup> | Int J Cancer, 1980 | All seven migrant source countries in 1970 had higher rates of stomach cancer than Australians, discussed with reference to inter-country dietary differences |
| Armstrong et al. <sup>53</sup> | Am J Epidemiol, 1983 | Mortality from Ischemic Heart Disease in Italians in Italy and Migrants 1/3 to 2/3 that of native-born Australians |
| Burvill et al. <sup>58</sup> | Soc Sci Med, 1983 | Suicide rates in migrants from England & Wales, Scotland and Ireland similar, although greater than in country of origin, especially Ireland |
| Mc Michael and Giles <sup>55</sup> | Cancer Research, 1988 | Review of cancers in migrants to Australia, suggesting strong evidence of environmental and behavioural influences |
| Mc Michael <sup>59</sup> | Community Health Studies, 1989 | Asserts that coronary heart disease is a mass disease of 20<sup>th</sup> century industrialised society requiring population-based prevention strategies |
| Hetzel et al. <sup>9</sup> | J Clin Epi, 1989 | Review of epidemiological and experimental studies related to fall in sudden deaths in USA and Australia since 1967 concludes that there is a protective effect of dietary polyunsaturates |
| Mc Michael et al. <sup>56</sup> | Med J Aust, 1989 | South Australia Cancer Registry data 1977-1986 show British and Irish migrants had higher incidence of all cancer sites combined, whereas Southern Europeans had lower rates |
| Ireland and Giles <sup>52</sup> | Aust NZ J Med, 1996 | Melbourne Collaborative Cohort Study 1990-1994, blood pressure higher in Italian-born middle-aged relative to Australian-born, not attributable to changes in diagnosis and treatment of hypertension |
| Burvill <sup>57</sup> | Psych Med, 1998 | Increased suicide rates in migrants relative to country of origin and importance of pre-migrant social and cultural experiences |

Italian and Greek immigrants to cities like Melbourne and Sydney initially had lower blood pressure and lipid profiles than their Australian born contemporary neighbours but had higher risk factor profiles than their home countries^50-53^. McMichael et al also reviewed cancer rates and there were differences in Northern European and Southern European immigrants^54-56^. Rates of stomach cancer, like stroke associated with early life experiences, tended to be higher in immigrant groups than native born Australians. Suicide rates were comparable in Northern European immigrants, but higher than in their country of origin^57-58^. The classical healthy migrant hypothesis posits that immigrants move from low to intermediate to high risk as they transition into a new environment^60^ though migrants usually form a relatively small part of the population, which is more typical of the later immigration patterns seen since the mid twentieth century in Australia. The post WW2 immigrants assume the risk of the prevailing environment as they enter middle and old age. McMichael was an early advocate of the general population drivers of cardiovascular diseases and supply factors underpinned by industrialisation^59^. There is contemporary interest in Australia in the health experiences of latter-day migrants, including in the Melbourne Collaborative Cohort Study^61^

It is likely that the effect of smoking is underestimated in these early epidemiological analyses. Active smoking in the post WW2 period was extraordinarily high, particularly in men^62^. Exposure to high levels of unrestricted active smoking would increase the risk of passive smoke exposure in public places and in households and transport. The current national population bans placing restrictions on public smoking in many countries have shown that admission rates for cardiovascular and respiratory disease are reduced in areas with effective smoking bans^63^.

### An Integrated Approach

The epidemiological focus has moved more recently to a more integrated lifecourse explanation for development of adult non communicable disease that takes account of early life exposures as well as personal health behaviours^6, 14, 21-23^. Policy should be directed at underlying drivers of population health as well as individual risk factor modification, as exemplified by current guidelines of management of hypertension^6^. Current rates of CSDs are much lower than in the past but show significant regional variations and inequalities and are now much higher in Aboriginal citizens ^64^.

Emerging contemporary data from other newer studies support the lifecourse hypothesis also^65-69^. The West Australian pregnancy cohort shows that adolescent adiposity trajectories are associated with increased risk of raised blood pressure^67^. Roberts and Wood employing data from a historical New Zealand cohort from 1907 to 1922 have modelled the significant effects of birthweight on subsequent adult blood pressure^68^. Taken together, these findings show the influence of lifecourse and the need also to understand the specific social and historical context.

In conclusion, this analysis provides a contextual reassessment of the historical drivers of the patterns of CSD in Australia. These different immigrant groups all experienced a period effect in relation to CSD, but also brought with them cohort effects of differing magnitude, which impacted CSD rates in the general population, reflected in the temporal association of age-adjusted CSD rates with immigration that later stabilised and in the age-year interactions in our models. With current global epidemics in developing economies exceeding in scale those in the past, these findings may be particularly relevant to public health policy planning for contemporary movements of peoples.

## Data Availability

All data produced in the present study are available upon reasonable request to the authors

## Author Contributions

CCK conceived the study design and wrote the first draft. JSMcC contributed the historical economic and public health context for Australia in the 19th and early 20th century. AMS and CCK conducted the literature search for contemporary Australia. JB initiated the construction of the database and RS conducted the temporal analysis. GEK devised and conducted the age period analysis for the general Australian and Foreign-born populations. All authors contributed to the interpretation of findings and to the drafting of the manuscript and agreed to its final content. Data files of these analyses may be provided, upon reasonable request, by the authors. There are no conflicts of interest to declare.

## Ethical Approval

This analysis employed publicly available archived databases with no individual participants so ethical approval was not sought or required.

## Funding and Acknowledgements

This analysis received no specific grant from any funding agency in the public, commercial or not-for-profit sectors. Professor Cecily Kelleher undertook the original research in Australia on which this paper is based with funding from a Miegunyah Fellowship at the University of Melbourne. Dr Jonathan Briody was funded by the Health Research Board SPHERE PhD training programme. Professor Janet McCalman is supported by the ARC, Australian Research Council. The authors wish to thank Ms Emma Coyle for the work on data entry for this analysis.

## Notes

### Competing Interest Statement

The authors have declared no competing interest.

### Funding Statement

This study did not receive any specific funding. Professor Cecily Kelleher undertook the research in Australia on which this paper is based with funding from a Miegunyah Fellowship at the University of Melbourne. Dr Jonathan Briody was funded by the Health Research Board SPHERE PhD training programme. Professor Janet McCalman is supported by the ARC, Australian Research Council.

### Author Declarations

Source data were openly available before initiation of the study. 1.Australian Bureau of Statistics. http://www.abs.gov.au/AUSSTATS/abs@.nsf/ViewContent?readform&view=ProductsbyCatalogue&Action=Expand&Num=2.2 accessed 12th January 2019 2.Australian Institute of Health and Welfare. https://www.aihw.gov.au/reports-data/health-conditions-disability-deaths/heart-stroke-vascular-diseases/overview, accessed 12th January 2019.

